# The Impact of Breast Cancer Polygenic Risk Score Disclosure on Decisional Conflict Around Risk-Reducing Mastectomy in *BRCA1/2* Carriers

**DOI:** 10.1101/2025.11.10.25339918

**Authors:** Giulia Ongaro, Caroline Salafia, Rania Sheikh, Elizabeth Schofield, Dana Farengo Clark, Jessica Ebrahimzadeh, Anu Chittenden, Jill E. Stopfer, Jamie Brower, Heather Symecko, Hannah Ovadia, Alexander Husband, Hadley Barr, Semanti Mukherjee, Jennifer L. Hay, Zsofia K. Stadler, Kenneth Offit, Judy E. Garber, Susan M. Domchek, Mark E. Robson, Jada G. Hamilton

## Abstract

**Purpose:** Female *BRCA1/2* carriers face high breast cancer risks and complex preventive decisions, which can generate decisional conflict. Polygenic Risk Scores (PRS) offer individualized risk estimates that may support decision-making. We examine whether PRS disclosure reduces decisional conflict in *BRCA1/2* carriers and whether PRS magnitude interacts with numeracy and tolerance for ambiguity.

**Methods:** Unaffected *BRCA1/2* carriers (*N=*354) were assessed pre PRS disclosure (T0), one-week (T1), and six-months post-disclosure (T2). PRS were translated into 10-year and lifetime absolute breast cancer risk estimates. Mixed-effects models tested changes in decisional conflict, and regression/moderation analyses tested PRS effects and interactions with numeracy and tolerance for ambiguity.

**Results:** Decisional conflict decreased significantly after PRS disclosure. Higher 10-year risk was associated with greater decisional conflict immediately after disclosure. Lifetime risk alone did not predict conflict. Tolerance for ambiguity moderated the effect of lifetime risk at T1: women with low/average tolerance experienced higher conflict with high PRS and lower conflict with low PRS. Numeracy, age, and family cancer history did not moderate PRS effects.

**Conclusion:** Decisional conflict decreased post-PRS disclosure, with short-term responses influenced by tolerance for ambiguity. Findings suggest that PRS can complement standard *BRCA1/2* genetic counseling and highlight the need for tailored risk communication.

## Introduction

Women with *BRCA1/2* pathogenic variants (PVs) (i.e., *BRCA1/2* carriers) face markedly increased lifetime risks of breast and ovarian cancer.^1^ For breast cancer in particular, cumulative risk estimates by age 80 vary widely—from 50%-79% for *BRCA1* carriers and 50%-77% for *BRCA2* carriers, compared to about 12% in the general female population—with risks reaching up to 85% among those with a particularly strong family cancer history.^2–4^ This variability reflects the incomplete penetrance of *BRCA1/2* PVs and influence of additional risk-modifying factors, including *BRCA* PV-specific risk, family cancer history, reproductive variables, lifestyle, environmental factors, and other common genetic variants.^1,5,6^

Considering this elevated and heterogeneous risk, *BRCA1/2* carriers must make complex and often emotionally charged decisions about breast cancer risk management, carefully weighing the benefits, harms, and level of protection offered by different strategies. Options for unaffected *BRCA1/2* carriers include enhanced surveillance (e.g., MRI, mammography), chemoprevention (e.g., tamoxifen), and risk-reducing surgery, such as risk-reducing mastectomy (RRM).^7^ Among these, RRM is the most effective, decreasing breast cancer risk by 90% or more.^8,9^ However, RRM involves irreversible physical changes and potential medical, psychological, social, and practical consequences.^10^ Complications may include infection, bleeding, chronic pain, loss of breast sensation, and revision surgeries, particularly with immediate breast reconstruction.^11^ Psychologically, women may struggle with changes in body image, sexual functioning, and identity.^12–14^ Surgical and recovery time can also disrupt personal, professional, and family life. For healthy and unaffected carriers—often identified through a relative’s cancer diagnosis and genetic testing—the choice can be especially challenging.^15^ As such, clinical guidelines stress that RRM decisions must be highly individualized and preference-sensitive, as evidence does not unequivocally support one option over another, and therefore, choices should be deliberated and informed.^16^

Given these complexities, many *BRCA1/2* carriers face substantial uncertainty when considering their options, with a sizeable proportion reporting meaningful levels of decisional conflict—a cognitive-emotional state of uncertainty about the best course of action, especially when choices involve complex trade-offs between life expectancy and quality of life.^15,17,18^ Specifically, about 43% of unaffected women with a *BRCA1/2* PV reported high decisional conflict when considering RRM.^17^ Various factors can shape the decision-making process and the decisional conflict it entails, including cognitive and psychological characteristics such as numeracy—the ability to understand and use numeric information—and tolerance for ambiguity—the capacity to manage uncertainty when outcomes are unclear—as well as demographic and experiential factors like age and family cancer history.^15,19–21^ Lower numeracy, greater intolerance of ambiguity, and younger age have been associated with more challenges in decision-making, and may contribute to higher decisional conflict, whereas women with a stronger family history may experience less uncertainty and conflict.^15,19,20,22^ Decisional conflict, in turn, has been linked to negative outcomes, including decision delay, decisional regret, and reduced satisfaction with chosen options.^23,24^ As such, understanding and mitigating decisional conflict is essential to improving decision-making quality in this context.

Recent advances in medical genetics have introduced polygenic risk scores (PRS) as a complementary approach to refine individual risk estimates.^2^ PRS condense into a single numerical value the small, additive effects of numerous common genetic variants (i.e., single nucleotide variants; SNV) associated with breast cancer risk in the general population and among *BRCA1/2* carriers.^25–29^ When incorporated into clinical risk models, PRS have the potential to stratify *BRCA1/2* carriers more precisely along a risk continuum, thereby clarifying the magnitude of risk, and informing individualized prevention and management strategies, while reducing uncertainty and supporting decisions.^30^ However, the complex, ambiguous, and probabilistic nature of PRS may also introduce new uncertainties, influencing decision-making processes, and potentially complicating rather than simplifying choices.^31^ It remains unclear whether integrating PRS into counseling and decision-making ultimately reduces decisional conflict for *BRCA1/2* carriers.

To establish the clinical and personal utility of PRS, it is important to understand how PRS-informed breast cancer risk estimates affect decisional conflict among *BRCA1/2* carriers. Addressing this gap is crucial as PRS are increasingly integrated into clinical practice. The present study evaluates the impact of PRS-informed breast cancer risk estimate disclosure (“PRS disclosure”) on decisional conflict regarding the choice between RRM and enhanced surveillance in unaffected *BRCA1/2* carriers, with two primary aims. First, to assess the effect of PRS disclosure on decisional conflict, we hypothesized a significant reduction in decisional conflict after PRS disclosure. Second, to assess whether the magnitude of PRS results is associated with decisional conflict, we hypothesized that individuals with higher PRS scores (i.e., greater breast cancer risk estimates) would experience lower decisional conflict. Additionally, we hypothesized that individual differences in numeracy, tolerance for ambiguity, age, and family cancer history would moderate this association.

## Materials and Methods

Eligible participants were English-speaking women ages ≥25 with a *BRCA1* or *BRCA2* PV and no personal history of breast cancer. Women who underwent RRM, had major psychiatric illness or significant cognitive deficits, or could not comply with study procedures were excluded. We recruited participants from three U.S. institutions: Memorial Sloan Kettering Cancer Center (MSK, New York, NY), the University of Pennsylvania (Penn, Philadelphia, PA), and Dana-Farber Cancer Institute (DFCI, Boston, MA). Potential participants were identified by their primary genetic counselor at the time of *BRCA1/2* PV disclosure or during routine follow-up appointments. At Penn, added outreach was conducted via recruitment letters and flyers to patients who had consented to future contact for research. Interested individuals received brief written educational information about PRS testing (described as “genetic risk modifier testing” to study participants^32^), provided informed consent, submitted a saliva sample for PRS testing, and completed the baseline assessment. Study measures were surveyed at three timepoints: baseline (T0), approximately one week after PRS disclosure (T1), and six months after PRS disclosure (T2). Participants received $10 in appreciation for completing each survey. All procedures were reviewed and approved by the Institutional Review Board. Recruitment lasted from April 2019-May 2024.

### PRS Testing

PRS were calculated based on validated algorithms^2,25^ leveraging 119 SNV previously associated with breast and ovarian cancer risk in the general population and in *BRCA1/2* carriers. Participants provided samples for genotyping using a clinical-grade testing platform (Phenogen Sciences); genotype reports were used to compute standardized PRS, which were subsequently translated into individualized risk estimates. Using R-package iCARE Individualized Coherent Absolute Risk Estimators, we built and evaluated models for absolute risk using SNV data and estimated an individual’s risk of developing disease during specific time intervals.^33^ Age-specific breast cancer incidence rates for *BRCA1* and *BRCA2* carriers were estimated from prospective cohort of *BRCA1* and *BRCA2* carriers.^2,30^ All participants received personalized PRS-informed breast cancer absolute risk estimates. Results were communicated to participants by trained genetic counselors using standardized visual risk communication materials developed and piloted at MSK.^32^ The risk communication materials (*Figure 1*) depicted reference values of the absolute remaining lifetime risk of developing breast cancer for the average woman of the same age (e.g., 12% for the average woman of age 40) and the range of absolute remaining lifetime risks of developing breast cancer for the average *BRCA1* or *BRCA2* carrier of the same age (e.g., 48%-78% for the average *BRCA1* carrier of age 40; 49%-81% for the average *BRCA2* carrier of age 40). Participants also received two distinct individualized risk estimates generated from their PRS values: (1) *absolute lifetime risk estimate*, which reflects an individual’s remaining lifetime risk of developing breast cancer accounting for their age, *BRCA* PV, and PRS, and (2) *10-year risk estimate*, representing the individual’s probability of developing breast cancer within the next 10 years accounting for their age, *BRCA* PV, and PRS.

**Figure 1.**
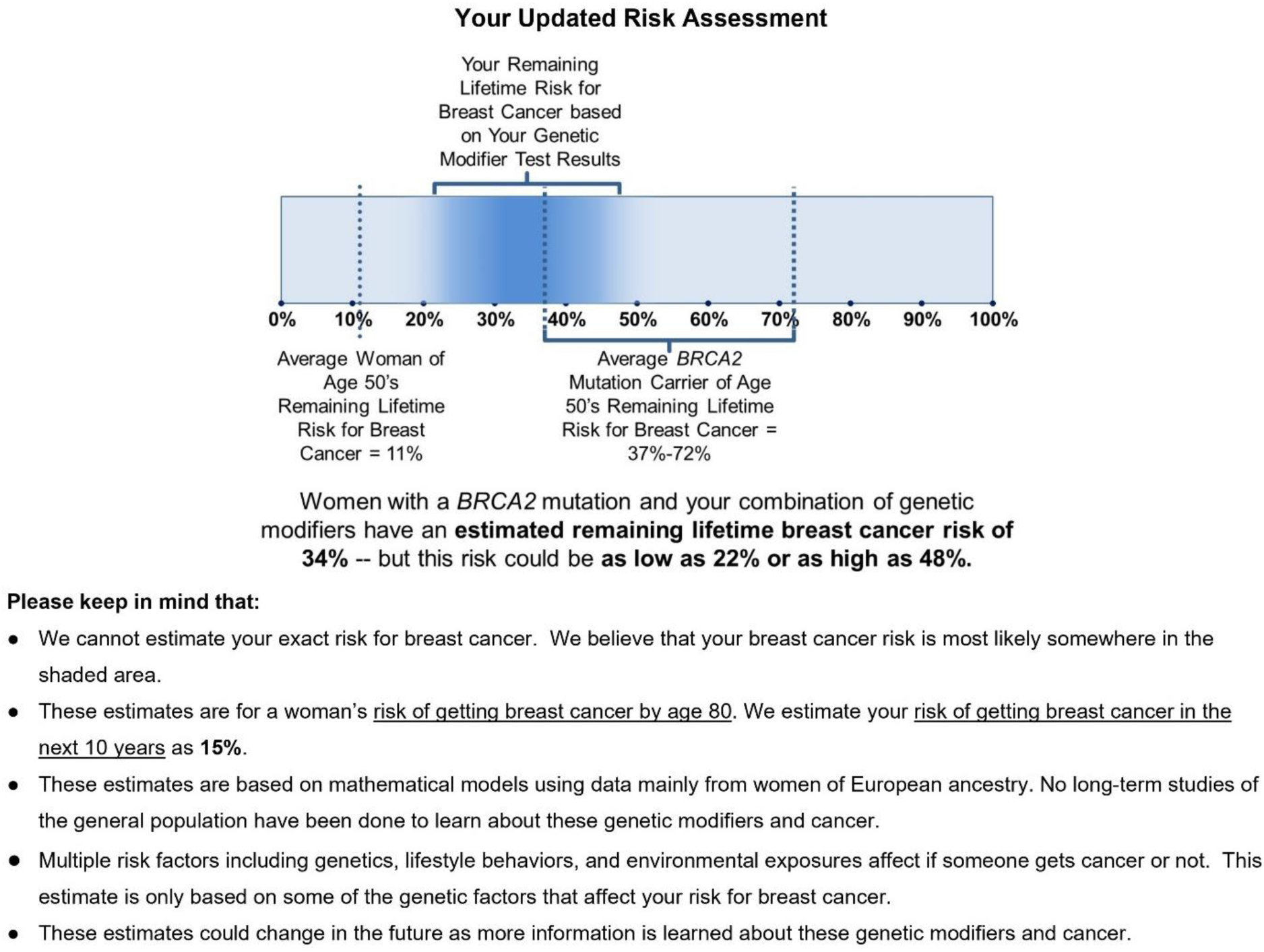
**Breast Cancer Risk Assessment: Example Report.**

### Measures

#### Sociodemographic and Clinical Characteristics

Age, race/ethnicity, income, education, parental status, insurance status, family history of any cancer and of breast cancer, and number of days since *BRCA* results disclosure were obtained from patient records when available or through self-report at T0.

#### Decisional Conflict

Decisional conflict regarding RRM was assessed using the Decisional Conflict Scale at all timepoints (DCS^23^). This instrument assesses perceptions of uncertainty when making healthcare decisions, contributing factors to uncertainty (e.g., feeling uninformed, unclear values, or unsupported), and perceived effectiveness of decision-making (i.e., feeling that the choice is informed, value-based, implementable, and satisfactory). Items were rated on a 5-point Likert scale, with total scores converted to a 0–100 scale, with higher scores indicating greater decisional conflict and analyzed as a continuous variable. Scores were also dichotomized based on two recommended cut-off scores^23^: scores 0-25 versus scores >25 indicated difficulty implementing decisions, and scores 0-37.5 versus scores >37.5 reflected clinically significant conflict often linked to delays or uncertainty. In this sample, the DCS showed excellent internal consistency (Cronbach’s α≥0.94).

#### Numeracy

Subjective numeracy was assessed at T0 using the Subjective Numeracy Scale,^34^ a self-report measure evaluating perceived mathematical ability and preference for numerical over verbal information. Responses were given on a 6-point Likert scale and averaged (range 1–6), with higher scores indicating greater subjective numeracy. The scale showed good internal consistency (α=0.85).

#### Tolerance for Ambiguity

Tolerance for ambiguity, defined as the capacity to manage uncertain, complex, or unfamiliar situations without experiencing distress, was assessed at T0 using the 7-item Tolerance for Ambiguity Scale.^35^ Participants rated their agreement with statements reflecting attitudes and emotional reactions to ambiguous circumstances on a 6-point Likert scale. Items were summed (range 7-42), with higher values indicating higher tolerance for ambiguity. This measure demonstrated acceptable internal consistency (α=0.76).

### Data Analysis

All variables were summarized using descriptive statistics. Data distribution and normality assumptions were assessed for continuous variables to determine the appropriateness of parametric tests. Potential differences related to study site were examined. Hierarchical linear models assessed changes in decisional conflict scores over time following PRS disclosure, adjusting for relevant covariates, including age, *BRCA* status, and family history of any and breast cancer (total number of first- and second-degree relatives with cancer). Participants who underwent RRM during follow-up did not provide subsequent DCS scores. To evaluate clinically meaningful changes in decisional conflict, we assessed the proportion of participants with high decisional conflict (scores >25 and >37.5) at each time point. We used a Cochran’s Q test for overall comparisons across time and McNemar’s tests for pairwise comparisons. Linear and logistic regressions examined associations between PRS result magnitude and decisional conflict, controlling for baseline values and relevant covariates (age, *BRCA* status, income, education). Moderation by numeracy, tolerance for ambiguity, age, and cancer family history were assessed via interaction terms. Analyses were two-tailed with α=0.05 and performed using SPSS version 29.

## Results

### Participants

A total of 354 women enrolled in the study (*M_age_*=39.1, *SD*=11.8, range: 25–79). The total of participants at each stage of the study is summarized in *Figure 2*. Response rates were 92.4% (T0), 77.1% (T1), and 70.6% (T2).

**Figure 2.**
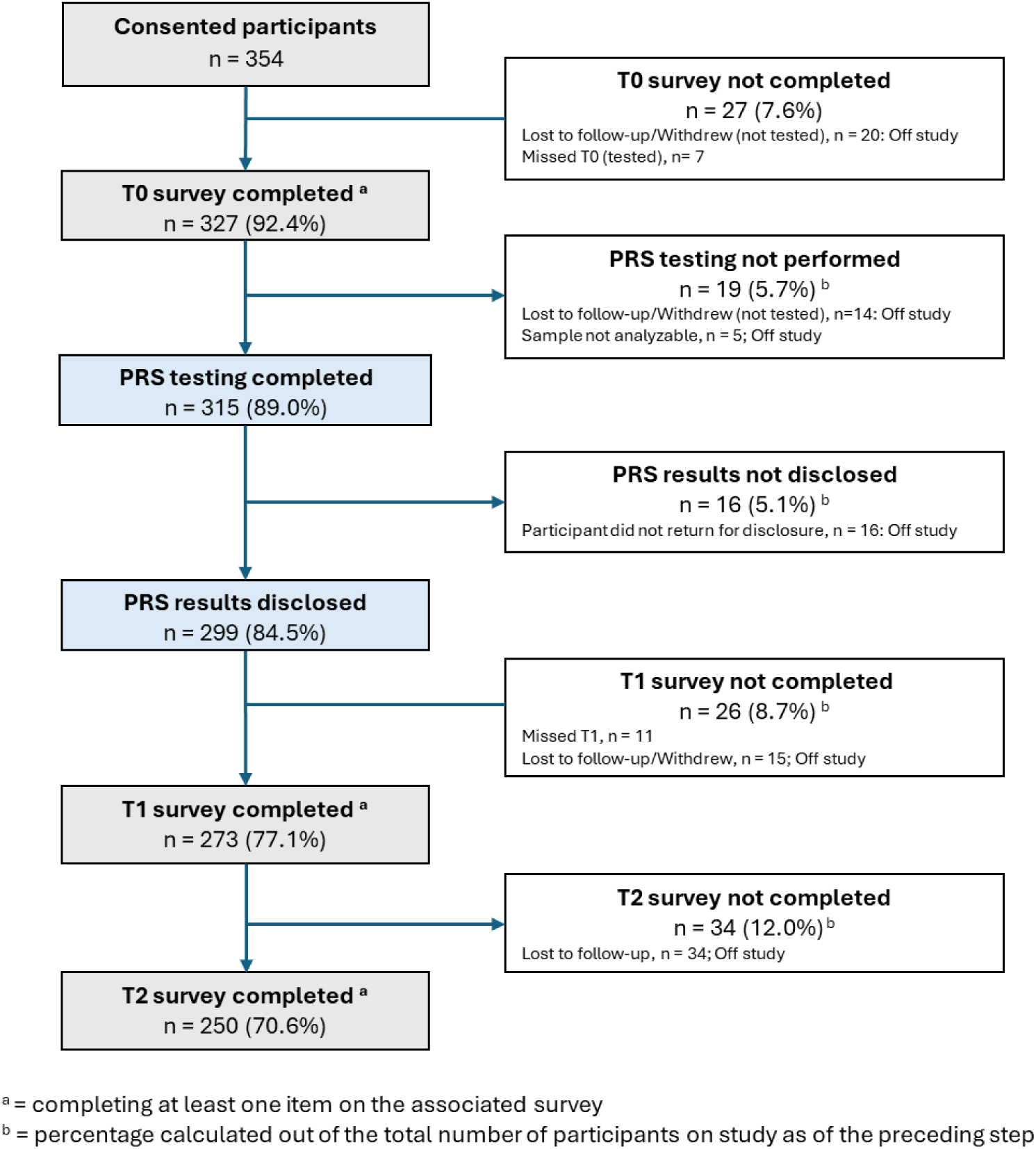
**CONSORT Flow Diagram.**

Most participants identified as White and non-Hispanic, were highly educated, with 97.5% having completed at least a college degree, and 64.1% reporting a household income of $100,000 or more. Most had private health insurance, and half had children. Among those with a family history of breast cancer, 31.1% reported the youngest age of diagnosis being under 40. The interval between *BRCA* results disclosure and baseline survey completion was available among those recruited from MSK and ranged from 0-4985 days (*M*=579.7, *SD*=987.7; *n*=108). Participants were comparable across sites in sociodemographic and clinical characteristics, except for age, with MSK participants being on average 5 years younger than those from Penn (*p*=0.004). Further descriptive details, including sample characteristics by study completion, are presented in *Table 1*.

**Table 1.**
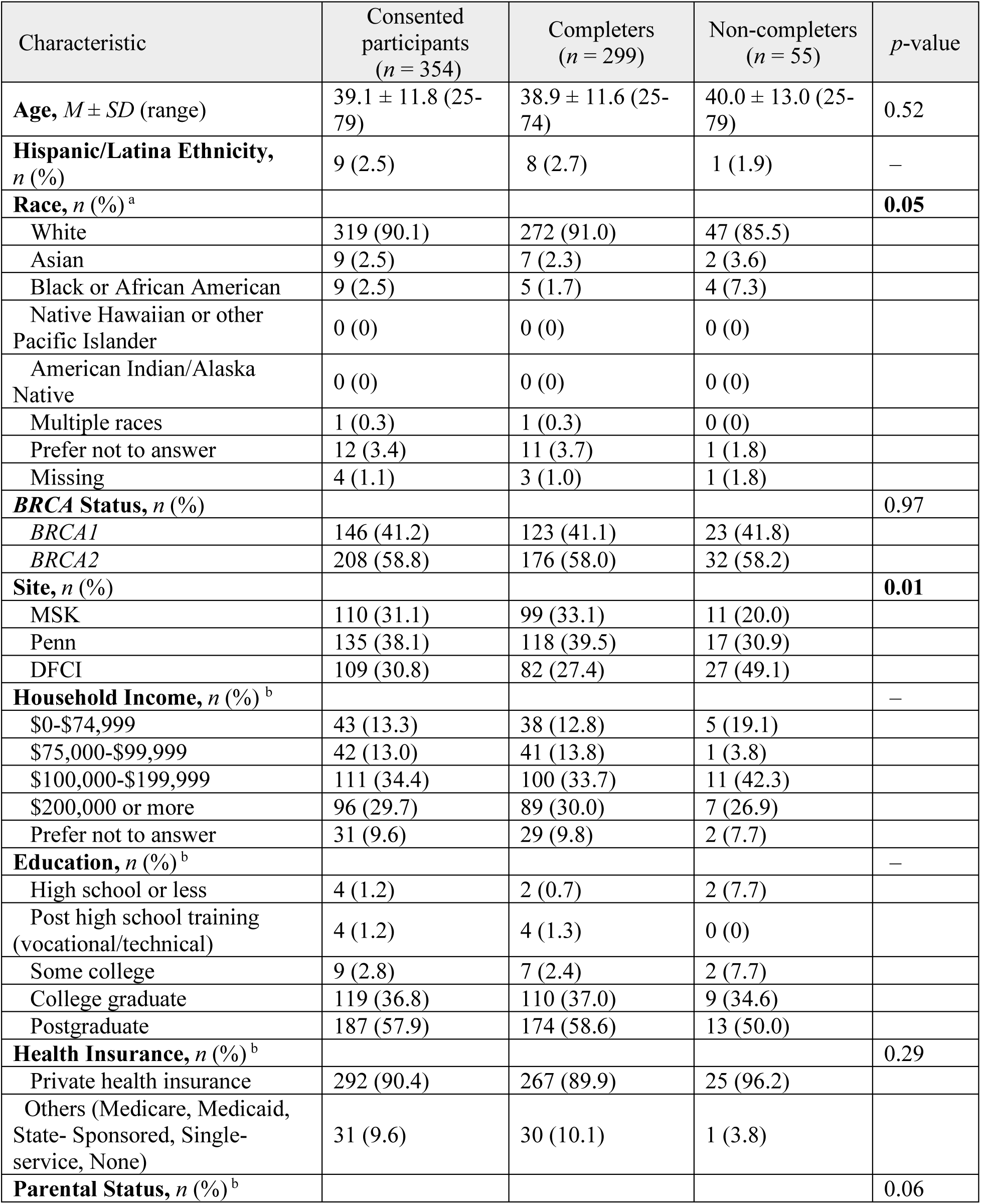
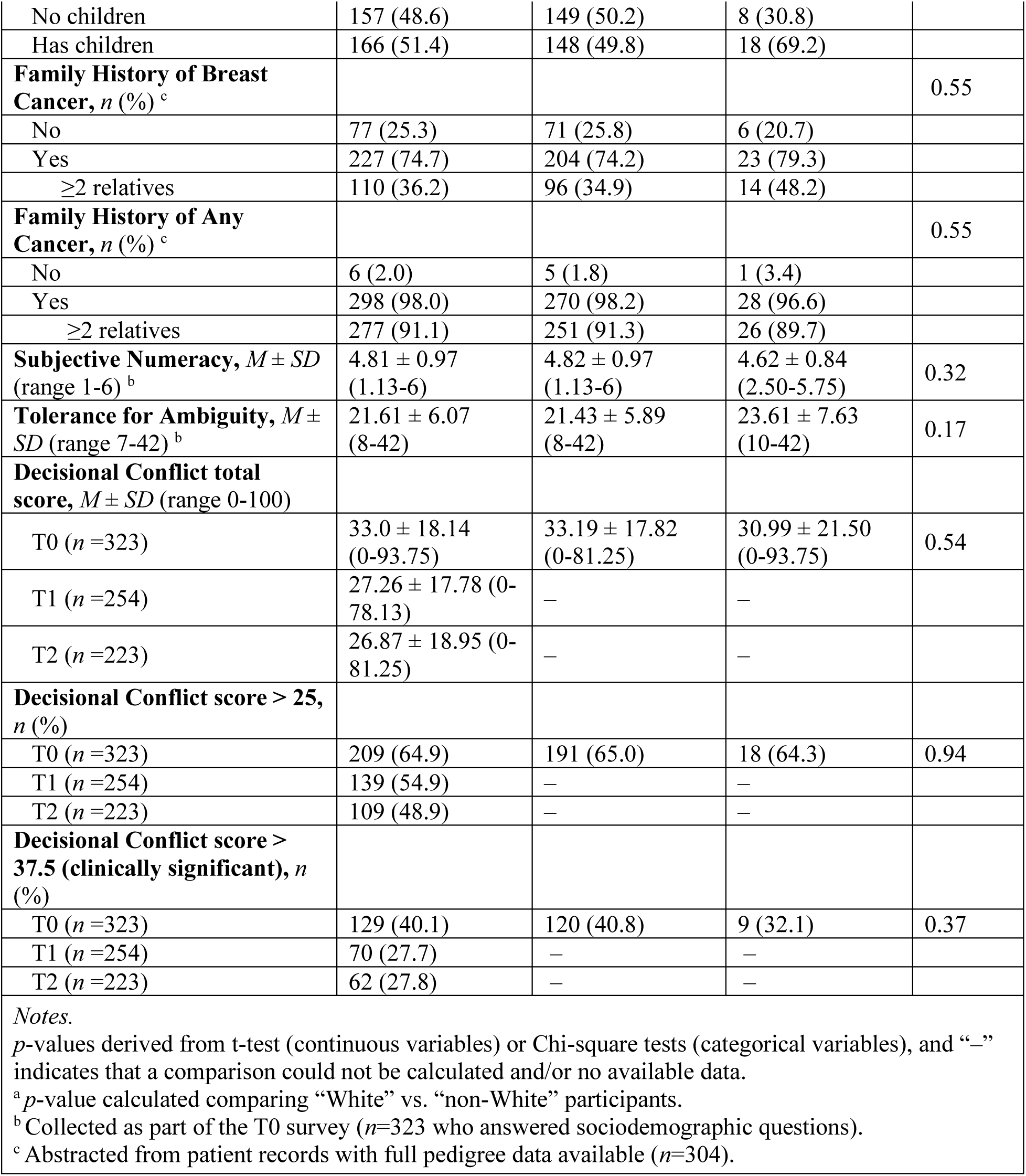
Sociodemographic and clinical characteristics of consented participants (*N* = 354) by study completion status, comparing completers and non-completers (i.e., those who did not complete the baseline T0 survey and/or PRS testing).

### Descriptive Statistics of Primary Measures

Descriptive statistics for numeracy, tolerance for ambiguity, and decisional conflict across all timepoints appear in *Table 1*. The table also reports the proportion of participants scoring above established cut-offs on the DCS, reflecting difficulty or clinically significant conflict in decision-making. Numeracy scores indicated generally high perceived numerical ability, while tolerance for ambiguity reflected moderate comfort with uncertainty. At T0, many participants reported considerable decisional conflict, with a substantial proportion exceeding thresholds for clinically meaningful conflict. Mean decisional conflict scores declined at T1 and T2; however, scores remained broadly distributed across the full-scale range. No significant correlations were observed between decisional conflict and time since *BRCA* results disclosure at any timepoint (all *p*>0.05).

PRS results were generated for 315 women in the study. As presented in *Figure 3*, the inclusion of PRS substantially shifted the distribution of participants’ estimated remaining lifetime breast cancer risks. Prior to PRS testing (i.e., based on age and *BRCA* PV; *Figure 3a*), estimates were tightly clustered around a narrow range, with a mean of 59.4% (*SD*=14.0). About two-thirds of participants (69.8%, *n*=220) had estimated risks between 60%-80%, and no participant had a risk estimate greater than 80%. When PRS was incorporated into the residual lifetime risk estimation (*Figure 3b*), although the mean remained similar (*M*=56.8%, *SD*=16.7), the distribution of estimated lifetime risks broadened considerably. Estimates ranged from 6%-97%, with 40% of the sample (*n*=126) falling between 60%-80%, reflecting a shift from the narrower distribution observed prior to PRS testing. Smaller proportions had estimates above 80% (6.4%, *n*=20) or below 20% (3.1%, *n*=10).

**Figure 3.**
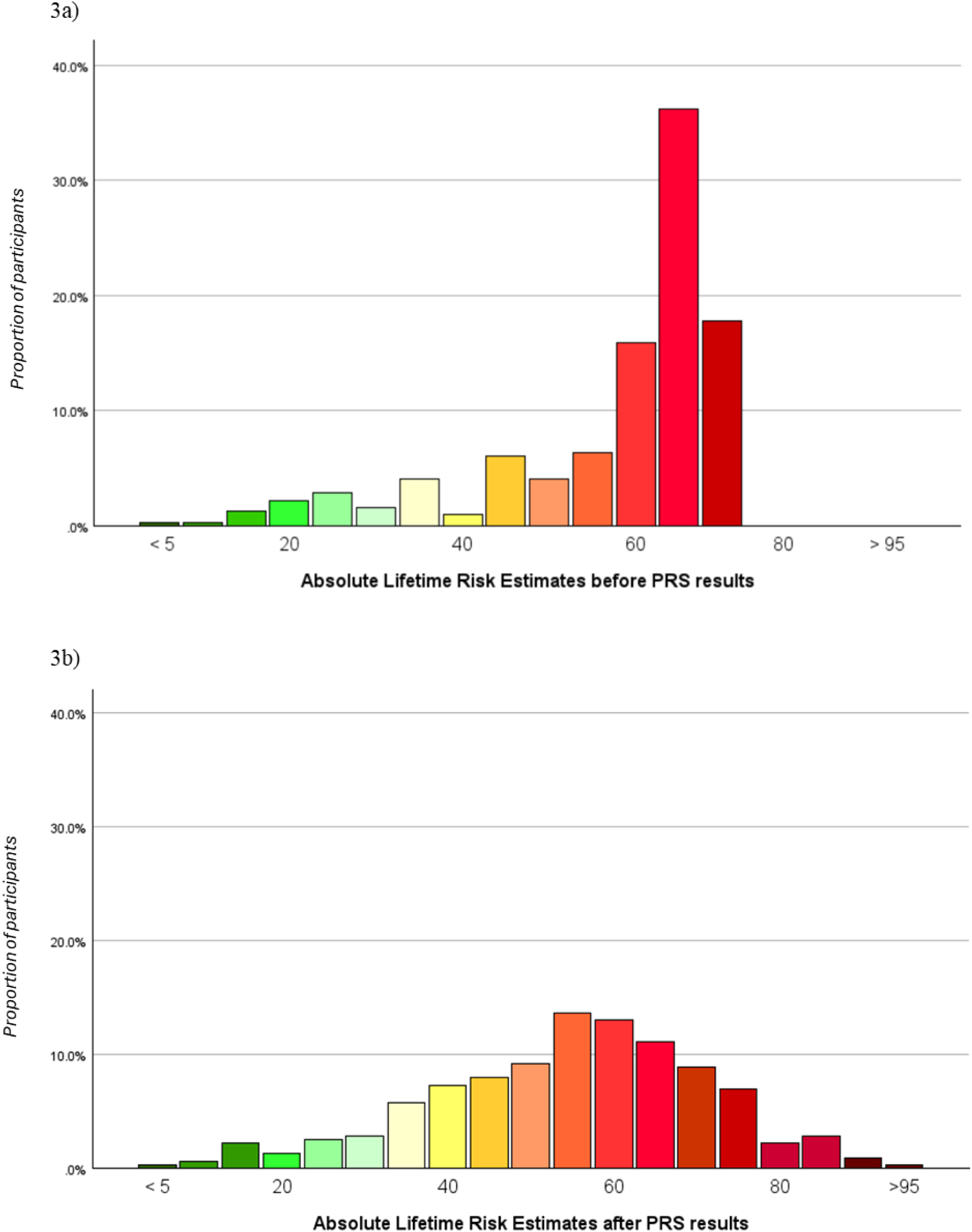
Distribution of breast cancer absolute remaining lifetime risk estimates before (a) and after (b) PRS results. *Notes*. Colors indicate risk levels, ranging from green for lower risk to red for higher risk. The gradient represents increasing absolute risk, with intermediate shades corresponding to intermediate risk levels.

### Change in Decisional Conflict after PRS Disclosure

To assess the impact of receiving PRS results on decision-making, we examined changes in decisional conflict over time among *BRCA1/2* carriers. Specifically, we tested whether DCS scores varied across the three timepoints. A linear mixed-effects model revealed a significant main effect of time on DCS scores (*F*(2, 505.04)=29.55, *p*<0.001). Compared to T0, DCS scores were significantly lower at both T1 (B= –6.10, *p*<0.001) and T2 (B= –6.37, *p*<0.001), indicating a significant and sustained reduction in decisional conflict following PRS disclosure. The fixed effect of time remained significant in a covariate-adjusted model that included age, *BRCA* status, and family cancer history (any or breast-specific) (T1: B= –6.43, *p*<0.001; T2: B= –6.30, *p*<0.001). No independent associations between covariates and decisional conflict were observed (*p*>0.16). The model had a marginal *R²*=0.045 and a conditional *R²*=0.673, suggesting most variance in decisional conflict was explained by between-participant differences.

To evaluate changes in clinically meaningful decisional conflict, we computed the proportion of participants exceeding established cut-off scores. Using the threshold of >25 (indicative of difficulty implementing a decision), 64.9% were above the cut-off at T0, 54.9% at T1, and 48.9% at T2. McNemar’s tests confirmed a significant reduction from baseline to post-disclosure (χ²(1)=10.72, *p*=0.001), with a non-significant downward trend from T1 to T2 (χ²(1)=3.51, *p*=0.061). Using the more stringent clinically significant threshold of >37.5 (associated with decision delay/uncertainty), the proportion above cut-off decreased from 40.1% at T0 to 27.7% at T1 and remained stable at T2 (27.8%), χ²(2)=23.12, *p*< .001.

Given the clinical relevance of sustained high decisional conflict, we conducted exploratory analyses among participants with clinically significant decisional conflict at T0 to identify characteristics associated with change. Specifically, we compared those whose decisional conflict remained high after PRS disclosure with those whose decisional conflict decreased below the clinically significant threshold at T1. No significant differences were found between the two groups in terms of sociodemographic factors (e.g., age, education), family cancer history (any or breast-specific), numeracy, or PRS results (i.e., absolute lifetime risk estimate, 10-year risk estimate) (*p*>0.22). The only factor that distinguished these groups was tolerance for ambiguity: women whose decisional conflict decreased showed significantly higher baseline tolerance compared with those whose conflict remained high (*M*=22.90 vs. 20.53; *p*=0.042; *d*=0.40), suggesting that greater tolerance for ambiguity may support a more adaptive psychological response to PRS-based risk information, even in the context of high-stakes decisions.

### Associations Between PRS Magnitude and Decisional Conflict

We examined whether the magnitude of PRS—expressed as either absolute lifetime risk or 10-year risk—predicted decisional conflict following PRS disclosure. Linear regression analyses were conducted separately at T1 and T2, first using unadjusted models (PRS only: lifetime risk or 10-year risk), followed by adjusted models controlling for baseline decisional conflict, age, *BRCA* status, income, and education (see *Supplemental Table 1*).

At T1, the unadjusted models indicated that higher lifetime risk estimates were significantly associated with greater decisional conflict (B=0.16, *p*=0.02), whereas 10-year risk estimates were not. After adjusting for baseline decisional conflict and covariates, the association with absolute lifetime risk was no longer significant; however, 10-year risk emerged as a significant predictor of decisional conflict (B=0.26, *p*=0.05). At T2, no significant associations were found between PRS (lifetime or 10-year) and decisional conflict. Across adjusted models, baseline decisional conflict was the strongest predictor, and in the T1 model including 10-year risk, older age was also associated with lower decisional conflict (B= –0.26, *p*=0.01). Model fit indices indicated acceptable fit across analyses.

Alternative models testing for potential curvilinear associations between PRS risk estimates and decisional conflict (i.e., both lower and higher PRS associated with reduced conflict) yielded no significant effects (all *p*>0.05).

### Moderation Analyses

Finally, we examined whether the relationship between PRS result magnitude and decisional conflict was moderated by individual-level characteristics, including numeracy, family cancer history (any or breast-specific), age, and tolerance for ambiguity. Separate multiple linear regression models were conducted for each proposed moderator and for each PRS result (absolute lifetime risk and 10-year risk), including an interaction term between PRS and the moderator, while adjusting for baseline decisional conflict, age, *BRCA* status, income, and education. No significant moderation effects were observed for numeracy, family cancer history (any or breast-specific), or age, for either PRS result (all *p*>0.10). In contrast, a significant moderating effect of tolerance for ambiguity was found only for the absolute lifetime risk estimate. The overall regression model was significant, *F*(8, 219)=21.28, *p*<0.001, explaining 42% of the variance in decisional conflict at T1 (Adjusted *R²*=0.42). The interaction between absolute lifetime risk and tolerance for ambiguity was significant (B= –0.02, *p*=0.033), indicating that the association between PRS magnitude and decisional conflict varied depending on participants’ ability to tolerate ambiguity. As shown in *Figure 4*, higher PRS-informed estimates of absolute lifetime risk were associated with greater decisional conflict—and lower scores with reduced decisional conflict—among women with low or average tolerance for ambiguity, while no such association was observed among those with high tolerance for ambiguity. This effect was no longer significant at T2 (*p*=0.40).

**Figure 4.**
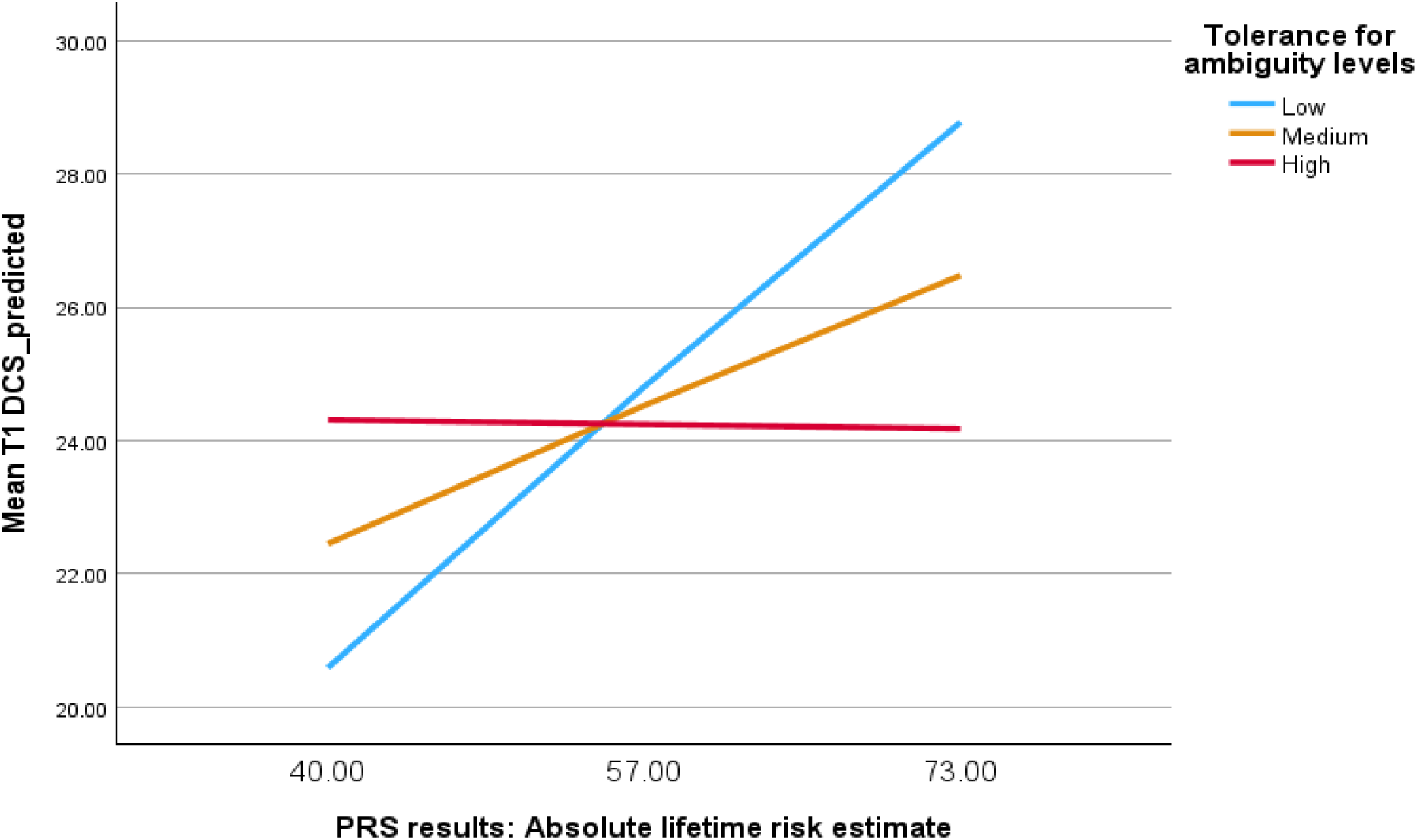
**Interaction between PRS Magnitude and Tolerance for Ambiguity in Predicting Decisional Conflict at T1. *Note.*** Predicted values of decisional conflict at T1 are plotted as a function of PRS and tolerance for ambiguity. In both cases, values were calculated at the mean (*M*) and ±1 standard deviation (*SD*). For tolerance for ambiguity, these are displayed as “low” (*M*–1 *SD*), “medium” (*M*), and “high” (*M*+1 *SD*); for PRS, numeric values are shown (40 = *M*–1 *SD*, 57 = *M*, 73 = *M*+1 *SD*).

## Discussion

We evaluated whether PRS disclosure affects decisional conflict among female *BRCA1/2* carriers—a population facing complex and high-stakes choices about breast cancer risk management. Consistent with our predictions, findings indicate a significant and sustained reduction in RRM decisional conflict over six months, both in mean scores and in the proportion of participants scoring above clinical cut-offs, suggesting that receiving individualized genomic risk information may help reduce uncertainty when considering preventive options.

Decisional conflict is dynamic and can be reduced through decision-support tools. In *BRCA1/2* carriers, approaches such as decision aids, shared decision-making, or specialized counseling can lower decisional conflict by providing information, clarifying options and personal values, and shaping patients’ risk perception.^18,36,37^ Within this framework, PRS disclosure—when delivered in the context of expert genetic counseling—may have contributed to reducing decisional conflict in our study by offering more individualized estimates of breast cancer risk, enhancing clarity, and helping patients feel better informed and more confident in their preventive decision-making. Prior work demonstrates that in high-risk women, disclosure of breast cancer PRS—particularly when accompanied by visual and narrative tools—can meaningfully shape risk perception without causing serious negative emotional effects.^38^ Other studies in high-risk populations without PVs, including cancer-affected and unaffected individuals, have demonstrated that receipt of higher PRS is associated with greater perceived breast cancer risk, shifting individuals’ understanding of their personal risk profile.^39,40^ PRS disclosure may clarify individual risk, thereby complementing standard genetic counseling with additional data that can be integrated into decision-making. However, caution is warranted in attributing these changes solely to PRS disclosure; although we confirmed that time since *BRCA* disclosure and DCS were unrelated within our sample, some reduction in decisional conflict could also reflect natural adaptation over time. Overall, our results indicate that decisional conflict is dynamic and that personalized genomic information, when integrated into counseling, may help women face uncertainty in breast cancer risk decisions.

Our findings also showed that the association between PRS magnitude and decisional conflict was nuanced and primarily observed in the short term. Ten-year risk estimates showed a significant association with decisional conflict immediately after disclosure such that women receiving higher 10-year risk estimates experienced greater decisional conflict and those receiving lower 10-year risk estimates experienced lesser conflict immediately after disclosure, while lifetime risk influenced decisional conflict only among women with lower tolerance for ambiguity. These findings indicate that the initial reaction to such personalized risk estimates can include momentary concern or uncertainty, particularly when perceived short-term risk is high. Furthermore, women who were less tolerant of ambiguity were highly reactive to this information, feeling reassured by low-risk scores but experiencing increased conflict with high-risk scores, whereas those with higher tolerance were relatively unaffected. This pattern resonates with prior work showing that individuals with low tolerance for ambiguity show stronger emotional and decisional reactions to probabilistic, ambiguous genetic risk information.^21^ Over time, enduring reductions in decisional conflict likely reflect a combination of factors, including baseline conflict, individual coping, and integration of the information within the context of expert counseling. These findings suggest that the extent to which PRS hold clinical or personal utility for *BRCA1/2* carriers varies based on the degree of predicted risk as well as on psychological traits. Thus, tailored communication and support that carefully considers these factors is likely valuable, maximizing the benefit of this information for women facing complex risk-management decisions.

This study has several strengths and limitations. Our relatively large sample, recruited across multiple U.S. institutions, enhances generalizability to *BRCA1/2* carriers in clinical care. However, participants were mostly non-Hispanic White, highly educated, and affluent, perhaps reflecting characteristics of individuals currently more likely to volunteer for PRS studies in high-risk contexts, while limiting generalizability to more socioeconomically and racially diverse populations. The applicability of PRS in non-European groups remains under investigation, and understanding its impact in diverse ancestral backgrounds is critical. Due to continued discovery of polygenes during the period of the planning of the project, we used a smaller SNP panel than currently available (i.e., 313 SNP panel ^27,29^) which may affect the generalizability of risk estimates. Additionally, PRS disclosure occurred in a specialized setting with expert counselors, and follow-up was short, leaving open questions about the relative effects of PRS versus counseling on decision-making, and whether PRS-related changes in decisional conflict influence uptake of preventive strategies. Finally, time since *BRCA* disclosure could only be accounted for in the MSK subset, reducing potential confounding, but future studies should further investigate its broader impact on decisional conflict. Nonetheless, PRS have promise for advancing precision prevention beyond high-risk settings, emphasizing the need for research in more representative cohorts, including underrepresented groups and varied healthcare contexts. Longer follow-up could clarify the durability of effects on decisional conflict and preventive behaviors, while understanding the psychological processes driving responses to PRS disclosure will be key to improving risk communication and decision support. Finally, prospective correlation of predictive psychosocial measures and endorsement of specific clinical interventions may allow more effective preventive cancer care.

Our study evaluated the impact of PRS-informed breast cancer risk estimates on RRM decisional conflict among unaffected *BRCA1/2* carriers. We observed a significant reduction in decisional conflict following PRS disclosure, with the effect moderated by psychological characteristics immediately post-disclosure, highlighting the need for tailored risk communication. These findings suggest that PRS can provide meaningful information to complement complex, preference-sensitive preventive decisions, enhancing the personal utility of genetic counseling. Together, these results underscore the promise of integrating PRS into clinical practice, while highlighting the need for future research to address diversity, long-term psychological and behavioral outcomes, including actual medical decisions, and optimal implementation strategies for different patient populations.

## Statements and Declarations

### Author Contributions

Conceptualization: G.O., K.O., J.E.G., S.M.D., M.E.R., J.G.H.; Formal analysis: G.O., E.S., J.G.H.; Funding acquisition: M.E.R., J.G.H.; Investigation: C.S., R.S., D.F.C., J.E., A.C., J.E.S., J.B., H.S., H.O., A.H., H.B., J.L.H., Z.K.S., J.G.H.; Methodology: S.M., K.O., M.E.R., J.G.H.; Project administration: J.B., A.H., H.O.; Writing – original draft: G.O., E.S., J.G.H.; Writing – review & editing: G.O., C.S., R.S., E.S., D.F.C., J.E., A.C., J.E.S., J.B., H.S., H.O., A.H., H.B., S.M., J.L.H., Z.K.S., K.O., J.E.G., S.M.D., M.E.R., J.G.H.

### Data Availability

The data presented in this study are available upon request from the corresponding author.

### Institutional Review Board Approval

The study has been approved by the Institutional Review Board of Memorial Sloan Kettering and all participating sites.

### Informed Consent

Informed consent was obtained from all participants involved in the study.

## Acknowledgements

The authors would like to thank the individuals who participated in this study, along with Philip Camille, Venecia Cano, Susan Holland, and Kaitlyn Lew for their contributions to participant recruitment and the execution of the study, and Erika Spaeth for engagement on behalf of the testing laboratory.

