## Supplemental Table 1 for "The Impact of Breast Cancer Polygenic Risk Score Disclosure on Decisional Conflict Around Risk-Reducing Mastectomy in *BRCA1/2* Carriers"

**Supplementary Table 1a-b. Linear Regression Coefficients Predicting Decisional Conflict at T1 and T2 from PRS Estimates.**

**1a.**

| **Predictor** | ***T1 – Decisional Conflict*** | | | | | ***T2 – Decisional Conflict*** | | | | |
| --- | --- | --- | --- | --- | --- | --- | --- | --- | --- | --- |
|  | **B** | **SE** | **Beta** | ***t*** | ***p*** | **B** | **SE** | **Beta** | ***t*** | ***p*** |
| **Unadjusted model** |  |  |  |  |  |  |  |  |  |  |
| PRS (Lifetime Risk) | 0.16 | 0.07 | 0.15 | 2.41 | 0.02 | 0.12 | 0.08 | 0.10 | 1.57 | 0.12 |
| Model Fit  (Adj. *R*²) |  |  |  |  | 0.019 |  |  |  |  | 0.006 |
| Model Fit (*F*(df)) |  |  |  |  | *F*(1,252) = 5.78 |  |  |  |  | *F*(1,222) = 2.44 |
| **Adjusted model** |  |  |  |  |  |  |  |  |  |  |
| PRS (Lifetime Risk) | 0.12 | 0.08 | 0.11 | 1.53 | 0.13 | 0.02 | 0.09 | 0.01 | 0.17 | 0.86 |
| T0- DCS | 0.61 | 0.05 | 0.63 | 12.10 | <0.001 | 0.66 | 0.06 | 0.63 | 11.16 | <0.001 |
| Age | –0.04 | 0.11 | –0.03 | –0.39 | 0.70 | –0.08 | 0.13 | –0.05 | –0.61 | 0.54 |
| *BRCA* status | 0.74 | 1.85 | 0.02 | 0.40 | 0.69 | –1.48 | 2.15 | –0.04 | –0.69 | 0.49 |
| Income ^a^ | –0.09 | 0.99 | –0.01 | –0.09 | 0.93 | 0.02 | 1.10 | 0.00 | 0.02 | 0.99 |
| Education ^b^ | 0.65 | 1.65 | 0.02 | 0.39 | 0.70 | 0.54 | 1.87 | 0.02 | 0.29 | 0.77 |
| Model Fit  (Adj. *R*²) |  |  |  |  | 0.41 |  |  |  |  | 0.38 |
| Model Fit (*F*(df)) |  |  |  |  | *F*(6,221) = 27.29 |  |  |  |  | *F*(6,194) = 21.81 |
| Note: ^a^ Income was modeled in four categories: $0–$74,999; $75,000–$99,999; $100,000–$199,999; ≥$200,000. ^b^ Education was modeled in three categories: some college or less; college graduate; postgraduate. | | | | | | | | | | |

**1b.**

| **Predictor** | ***T1 – Decisional Conflict*** | | | | | ***T2 – Decisional Conflict*** | | | | |
| --- | --- | --- | --- | --- | --- | --- | --- | --- | --- | --- |
|  | **B** | **SE** | **Beta** | ***t*** | ***p*** | **B** | **SE** | **Beta** | **t** | **p** |
| **Unadjusted model** |  |  |  |  |  |  |  |  |  |  |
| PRS (10-year Risk) | 0.19 | 0.14 | 0.09 | 1.40 | 0.16 | 0.08 | 0.15 | 0.04 | 0.55 | 0.59 |
| Model Fit  (Adj. *R*²) |  |  |  |  | 0.004 |  |  |  |  | -0.003 |
| Model Fit (*F*(df)) |  |  |  |  | *F*(1,251) =1.95 |  |  |  |  | *F*(1,222) =0.30 |
| **Adjusted models** |  |  |  |  |  |  |  |  |  |  |
| PRS (10-year Risk) | 0.26 | 0.13 | 0.12 | 1.97 | 0.05 | 0.06 | 0.15 | 0.03 | 0.43 | 0.67 |
| T0- DCS | 0.61 | 0.05 | 0.63 | 12.04 | <0.001 | 0.66 | 0.06 | 0.62 | 11.14 | <0.001 |
| Age | –0.26 | 0.10 | –0.16 | -2.65 | 0.01 | –0.12 | 0.11 | –0.07 | -1.05 | 0.30 |
| *BRCA* Status | 1.91 | 1.97 | 0.05 | 0.97 | 0.33 | –1.19 | 2.27 | –0.03 | -0.52 | 0.60 |
| Income ^a^ | –0.23 | 0.99 | –0.01 | -0.23 | 0.82 | –0.04 | 1.11 | –0.00 | -0.04 | 0.97 |
| Education ^b^ | 0.70 | 1.67 | 0.02 | 0.42 | 0.68 | 0.53 | 1.87 | 0.02 | 0.29 | 0.78 |
| Model Fit  (Adj. *R*²) |  |  |  |  | 0.42 |  |  |  |  | 0.39 |
| Model Fit (*F*(df)) |  |  |  |  | *F*(6,220) = 27.71 |  |  |  |  | *F*(6,194) = 21.85 |
| Note: ^a^ Income was modeled in four categories: $0–$74,999; $75,000–$99,999; $100,000–$199,999; ≥$200,000. ^b^ Education was modeled in three categories: some college or less; college graduate; postgraduate. | | | | | | | | | | |
